# Sex differences in the change in cardiorespiratory fitness and additional physical and mental health outcomes after exercise training in adults with atrial fibrillation: a systematic review protocol

**DOI:** 10.1101/2022.05.19.22275338

**Authors:** Sol Vidal-Almela, Isabela R. Marçal, Tasuku Terada, Carley D. O’Neill, Jennifer L. Reed

## Abstract

**Background:** Patients with atrial fibrillation (AF), the most common sustained cardiac arrhythmia, often have a low cardiorespiratory fitness (CRF) and poor physical and mental health due to disabling AF symptoms. This is more pronounced in females, who also report worse AF symptoms and quality of life (QoL) than males. Improving CRF through exercise training is an important AF management target associated with lower hospitalization and mortality rates. Emerging research suggests smaller CRF improvements in females than males following the same exercise training program. Yet, this has not been systematically reviewed in the AF population. The primary purpose of this systematic review is to compare changes in CRF following exercise training between females and males with AF. Secondary aims will compare changes in AF symptoms, QoL and additional physical and mental health outcomes between sexes.

**Methods:** We will adhere to the reporting guidelines of the Preferred Reporting Items for Systematic Review and Meta-Analyses (PRISMA) statement. Five electronic bibliographic databases are being searched to identify studies with prospective cohort and experimental designs, implementing exercise training of any form (e.g. aerobic, strength) for at least 4 weeks, in adults (≥18 years old) with an AF diagnosis. Eligible studies must report a baseline and follow-up measure of at least one primary or secondary outcome. CRF (primary outcome) can be estimated or directly measured as peak oxygen consumption (VO_2_peak). When eligible results are not segregated by sex, authors will be contacted to obtain sex-specific data. Study quality and risk of bias will be assessed using the Tool for the assEssment of Study qualiTy and reporting in EXercise (TESTEX) scale and the Grading of Recommendations Assessment, Development and Evaluation (GRADE) approach. Meta-analyses will be conducted to synthesize the measures of effect in studies with sufficient homogeneity.

**Discussion:** This review will address the lack of sex-based analyses in exercise studies in the AF population. By using a sex lens, we will provide evidence on the physical and mental health effects of exercise training in females and males with AF. Our findings will be of value to patients with AF, researchers and healthcare providers involved in AF management.

**Systematic review registration:** PROSPERO #CRD42022302310

## BACKGROUND

Atrial Fibrillation (AF) is characterized by an irregular cardiac rhythm and rapid heart rate. There are currently more than 37 million patients with AF globally (1,2), and a 60% increase in AF prevalence is expected by 2050 (2). AF is typically a progressive disorder (paroxysmal to persistent and finally permanent) due to the accumulation of modifiable AF substrates (e.g. hypertension, obesity) (3,4), yet participation in exercise-based cardiovascular rehabilitation has been shown to lower the likelihood of AF progression (5).

Several research trials implementing exercise training have reported improvements in cardiorespiratory fitness (CRF) in patients with AF (6–16). CRF independently predicts AF recurrence (17) and prognosis (18,19); every 1 metabolic equivalent (MET; i.e. 3.5 mL O_2_·min^−1^·kg^−1^) increase in CRF reduces AF recurrence by 9% (17). Patients with AF have a 3.3 to 5.0 mL O_2_·min^−1^·kg^−1^ lower CRF than other cardiovascular disease populations (11,16) and are bothered by a low exercise tolerance (20). An elevated left ventricular filling pressure and diminished chronotropic response contribute to the reduced CRF in AF (21). In these patients, improving CRF with exercise-based cardiovascular rehabilitation is associated with a lower risk for cardiovascular disease hospitalization or all-cause mortality (hazards ratio 0.83, 95% confidence interval [CI] 0.69 to 0.98, p=0.04) (22).

Females with AF have a substantially lower CRF than males (23). Females’ lower CRF and physical activity levels (24) may be associated with their reporting of more frequent and severe AF symptoms (e.g. dyspnea, fatigue, palpitations, dizziness, anxiety) (25–27); poorer quality of life (QoL) (28,29); and greater risk of mortality when compared to males (2). Improving CRF through exercise training represents, thus, an important AF management target which may be particularly beneficial for females.

Some studies in healthy and cardiovascular disease populations suggest that females may improve CRF to a lesser extent than males following the same exercise program (30–34). In the CopenHeartRFA trial (n=59 females, 151 males), females with paroxysmal or persistent AF experienced a smaller increase in CRF than males (+1.7 vs. +2.9 mL·min^−1^·kg^−1^) following 12-weeks of combined aerobic and strength training (23). Similarly, a 12-month walking intervention in patients with permanent AF (n=7 females, 13 males) led to an increase in CRF of 7% in females and 18% in males (11). Since only two studies in the AF population have reported CRF segregated by sex, additional comparisons of CRF changes following exercise training between females and males are warranted by pooling sex-specific data during meta-analyses (35).

Exercise training has also been shown to improve AF symptoms (10,36) and QoL (8,10,12,14,36) in patients with AF. The CopenHeartRFA trial reported significant increases in AF-specific QoL as measured by the atrial fibrillation effect on quality-of-life (AFEQT) scale in females (global score: +34.4±16.9 points; treatment concern score: +17.0±23.5 points) but not males, when compared to usual care (23). Exercise may narrow pre-existing differences in QoL between sexes.

There are sex-based differences in additional modifiable risk factors for AF. The prevalence of hypertension (27,28,37), obesity (24) and depression (24) is higher in females than males with AF. Females also experience faster heart rates during AF (38), which may accelerate the electrical and structural remodeling in the atria (39). Such risk factors contribute to AF pathogenesis (39), but irrefutable evidence has shown the benefits of exercise training in improving physical and mental health (40,41). In patients with AF, whilst exercise alone may not improve blood pressure (6,12,16,42,43), it has been shown to reduce body mass (10) and fat mass (11,12), anxiety and depressive symptoms (8,42), and resting heart rate (7,10,12,13,16). Less is known about the sex-based differences in these outcomes following exercise training in patients with AF (23). In other cardiovascular disease populations, findings have been inconsistent (32–34,44,45). A recent meta-analysis in patients with coronary artery disease revealed no sex-based differences in blood pressure reductions (n=6 studies, systolic: between sexes mean difference [MD] -2.51, 95% CI -6.57 to 1.55 mmHg, p=0.23; diastolic: MD -0.91, 95% CI -4.98 to 3.15 mmHg, p=0.66) and lower improvements in body mass index (BMI) (n=10 studies, MD -0.25, 95% CI -0.03 to -0.47 kg/m^2^, p=0.02) in females than males following high-intensity interval training (45).

Exercise training is now part of AF care (4,46,47), yet research gaps remain regarding sex-based differences in the response to exercise training in the AF population (48). A systematic evaluation and synthesis using a sex lens is needed as a first step to provide evidence-based exercise recommendations for patients with AF applicable to both sexes.

### Objective

The primary purpose of this systematic review is to compare changes in CRF following exercise training between females and males with AF. We hypothesize that across available studies, smaller improvements in CRF will be reported following exercise training for females with AF than males. The secondary aim is to examine the impact of sex on changes in AF symptoms, general and AF-specific QoL, and additional physical and mental health outcomes (e.g. resting blood pressure and heart rate, BMI, body composition, functional capacity, physical activity levels, anxiety and depressive symptoms) following exercise training.

## METHODS

### Study design

This systematic review protocol was designed in accordance with the Preferred Reporting Items for Systematic Review and Meta-Analyses (PRISMA)-P checklist (49) (see Additional File 1). The systematic review will adhere to the most updated reporting guidelines of the PRISMA statement (50) and will follow the criteria outlined in the checklist A Measurement Tool to Assess Systematic Reviews (<u>AMSTAR</u>) (51).

### Study registration

This systematic review protocol was registered with the International Prospective Register of Systematic Reviews (PROSPERO) in February 2022 (registration # CRD42022302310). In the event of important protocol amendments, the date of each amendment accompanied by a description of the change and the rationale will be reported in PROSPERO.

### Eligibility criteria

#### Participants

Studies will be included if the sample is comprised of adults (≥18 years old) with an AF diagnosis and where data from adults with AF can be extracted (i.e. ≥10% of the sample has AF). There will be no restrictions in co-morbidities.

#### Study designs

Prospective experimental designs (randomized controlled trials [RCTs], quasi-experimental, pre-post, and case-control studies) will be eligible.

#### Interventions

Eligible studies must contain an intervention component incorporating exercise training of any form with a duration of at least 4 weeks (52). We will use the well-established definition by Caspersen et al, where exercise is defined as a structured, planned and repetitive subset of physical activity with the aim to maintain or improve physical fitness and health (53). The exercise interventions may include, but are not limited to, aerobic training (e.g. moderate-intensity continuous training, high-intensity interval training, circuit training), strength training (e.g. body-weight or machine-based resistance training, isometric exercises) and Yoga. The interventions may be multi-component, but they must include exercise. There will be no restrictions on the mode of delivery (e.g. in person, virtual, hybrid) or setting (e.g. in hospital, in the community, individual or as a group) of the intervention.

#### Outcomes

The primary outcome of interest will be CRF. In clinical populations, CRF is often measured as the highest value of oxygen consumption (VO_2_peak) attained during an exercise test designed to bring the participant to volitional fatigue (54), or estimated from a submaximal exercise test (55). Thus, CRF will be considered as directly measured or estimated VO_2_peak (in mL·min^−1^·kg^−1^, L·min^−1^ or METs). Self-reported measures of VO_2_peak using a questionnaire (e.g. Duke Activity Status Index) will be excluded.

Eligible studies must report a baseline and follow-up measure of at least one primary or secondary outcome. Secondary outcomes will consist of:

#### AF symptom frequency and/or severity

(patient-reported outcome) measured with self-report questionnaires which may include, but are not limited to, the University of Toronto Atrial Fibrillation Severity Scale (AFSS), European Heart Rhythm Association (EHRA) AF symptom scale, Arrhythmia Symptom Checklist Frequency and Severity (SCL), Canadian Cardiovascular Society Severity in Atrial Fibrillation Scale (CCS-SAF), Mayo Atrial Fibrillation Symptom Inventory (MAFSI), and Arrhythmia-Specific Questionnaire in Tachycardia and Arrhythmia (ASTA) (56,57).

#### General QoL

(patient-reported outcome) measured with self-report questionnaires which may include the Medical Outcome Study Short-form Health Survey (SF-12 or SF-36) and EuroQoL (EQ-5D) (56).

#### AF-specific QoL

measured with self-report questionnaires such as the AFEQT, Atrial Fibrillation Quality of Life (AF-QoL), Quality of Life in Atrial Fibrillation (QLAF) and Atrial Fibrillation Quality of Life Questionnaire (AFQLQ) (56).

#### Additional physical health outcomes

(i.e. resting blood pressure and heart rate, BMI, and body composition) measured by research or clinical staff. Functional capacity measured with a 6-minute walking test. Physical activity levels measured by a self-report questionnaire or a device such as an accelerometer.

#### Mental health outcomes

(patient-reported outcome) including anxiety and depression levels, measured with self-report questionnaires which may include, but are not limited to, Hospital Anxiety and Depression Scale (HADS), Generalized Anxiety Disorder (GAD-7), Beck Anxiety Inventory (BAI), State-Trait Anxiety Inventory (STAI), Patient Health Questionnaire-9 (PHQ-9), Beck Depression Inventory (BDI), and Cardiac Depression Scale (CDS).

Additional outcomes will be compared between sexes including: (a) adherence as defined by the mean number of sessions attended by participants, (b) number of dropouts and loss-to-follow-up; and (c) number of adverse events during the study period.

#### Publication status and language

Published peer-reviewed journal articles and non-peer reviewed grey literature including published theses and dissertations (e.g. ProQuest Dissertations & Theses Global, GreyLit.org, Open Grey), conference abstracts, presentations and proceedings; and unpublished data will be examined. No language restrictions will be imposed on the search, but only papers written in English, French, Japanese, Portuguese or Spanish will be included.

### Search strategy

A comprehensive search strategy from an existing systematic review by the authors (58) was re-used to identify relevant studies. This search was previously designed and translated according to the indexing systems of several databases in collaboration with a medical research librarian. The search was previously peer reviewed by a second research librarian. Medical subject headings (MeSH) and key words related to “atrial fibrillation”, “exercise training” and “cardiac rehabilitation” were used. A draft of the search strategy used for MEDLINE is provided in Table 1. No date restrictions were imposed. Five electronic bibliographic databases were searched: MEDLINE (OvidSP); CINAHL (EBSCOhost); Embase (OvidSP); PycINFO (Ovid) and Cochrane Library (Wiley). The reference lists of studies selected for the review and those of previous reviews will be examined. Registries such as Clinical Trials and PROSPERO will be searched for recently completed trials and systematic reviews, respectively; and conference proceedings of the two previous years related to the AF and exercise field will be hand searched to identify further studies.

**Table 1.**
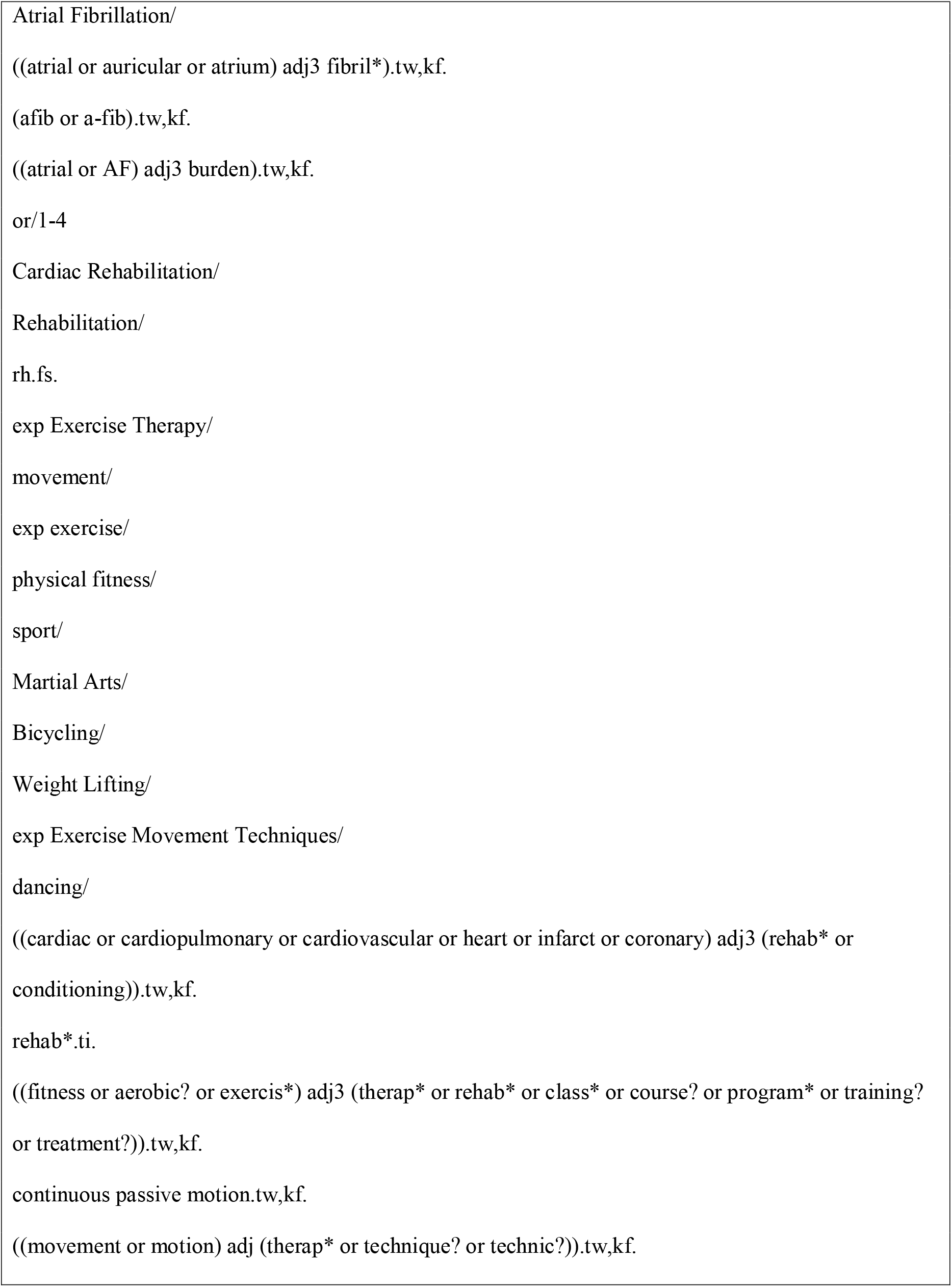
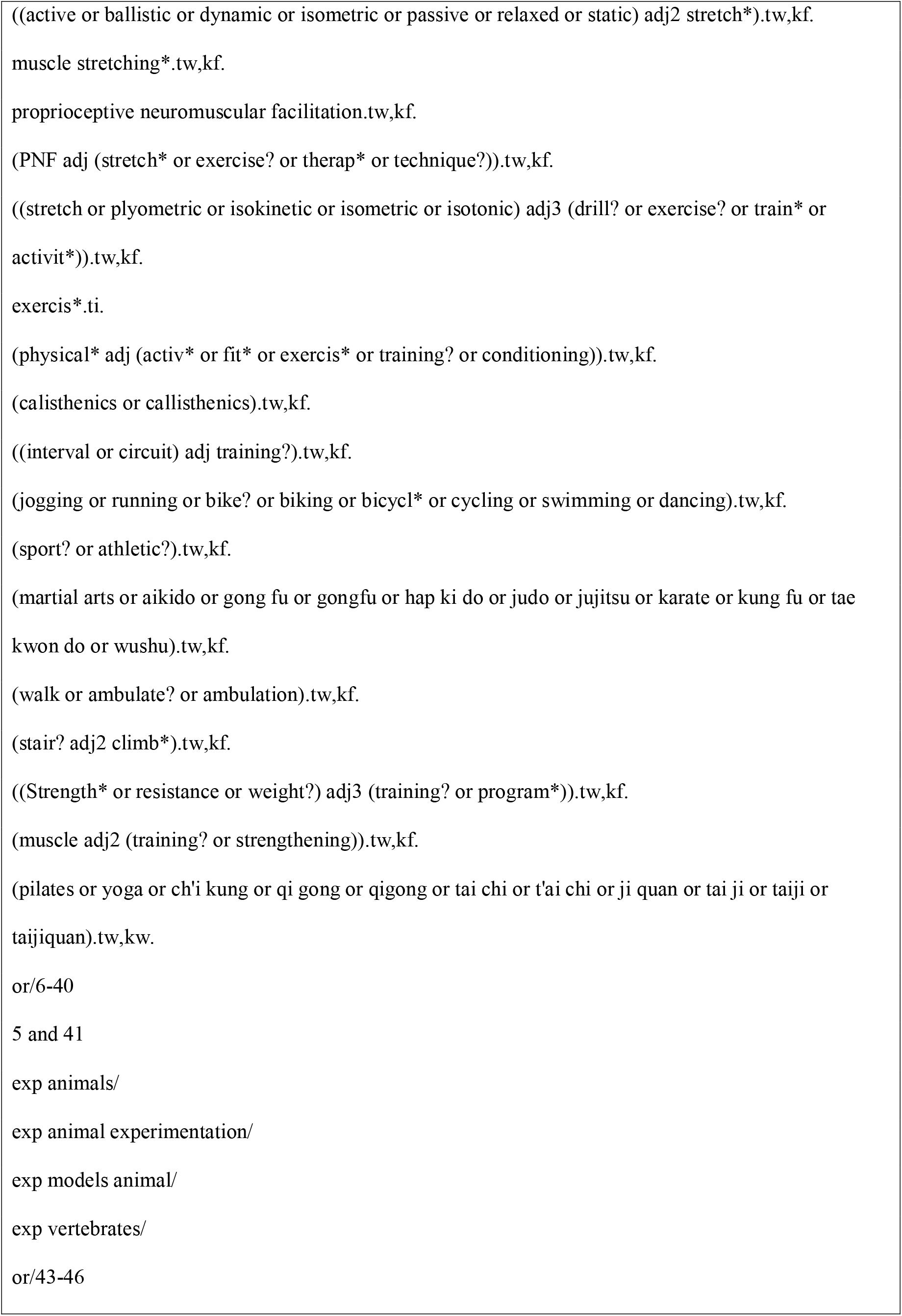
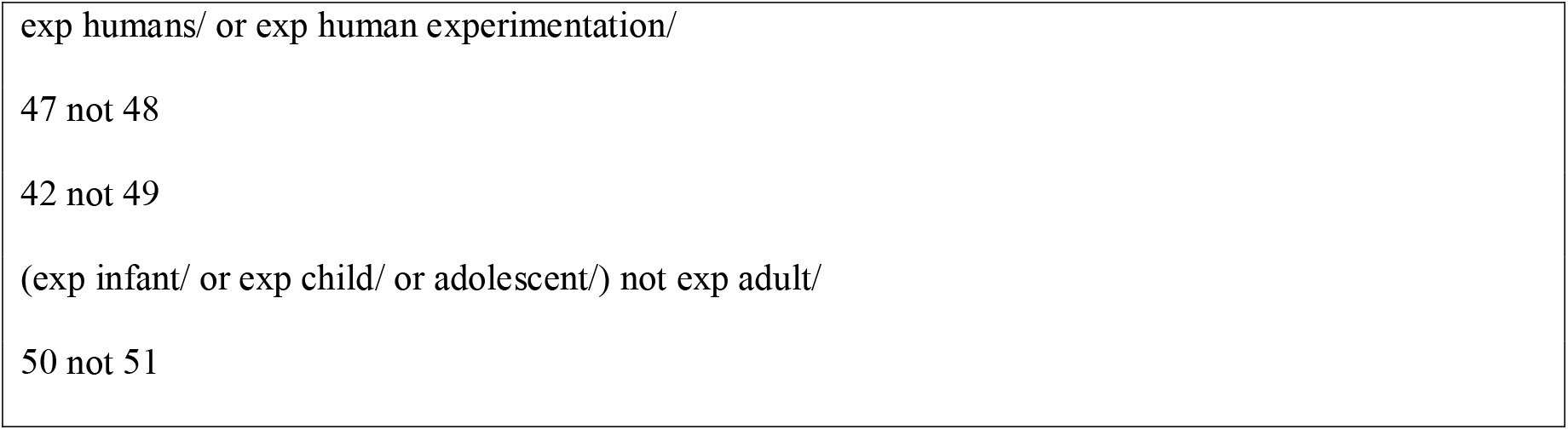
Example of the search strategy for MEDLINE.

### Data management and selection of eligible studies

The results from the search strategy have been uploaded into Covidence Systematic Review Software (Veritas Health Innovation, Melbourne, Australia), an electronic program that facilitates collaboration among reviewers during the study selection process. Duplicate articles were removed using the “duplicate” function and remaining duplicates will be excluded manually by two reviewers during the screening process (SVA, IRM). If several articles are published using the same data source, the articles with the largest sample size for CRF, with additional secondary outcomes, or reporting sex-specific results may be retained. Labels with specific reasons for exclusion have been created in Covidence to facilitate the screening process and the creation of a flow chart. Two independent reviewers (SVA, IRM) are screening the titles and abstracts of all articles yielded by the search against the inclusion criteria. Prior to formally beginning screening, a calibration exercise was conducted. Full texts of each potentially relevant article will be uploaded on Covidence and screened by two independent reviewers (SVA, IRM) to determine eligibility. Any disagreements between the reviewers or any uncertainty will be resolved by discussion with a third reviewer (JLR). The review authors will not be blinded to the study authors or journal titles when screening.

### Data extraction

A data extraction form has been created using Microsoft Excel (Microsoft Canada Inc. Mississauga, ON, Canada) (see Additional File 2) and will be adapted following feedback from co-authors to improve its usability. Data extracted from each eligible study, when available, will include: publication details (authors, country and year of publication), participant characteristics by sex (sample size, AF type, mean age, sociodemographic and gender-related variables [e.g. gender identity, ethnicity, marital status, educational level, employment status, income], prescribed medications, medical conditions and procedures), study design, characteristics of the assessment of VO_2_peak (test protocol, modality, method of measurement), details of exercise interventions (frequency, intensity, time, type, [FITT], volume and progression), participant adherence with the exercise intervention (number of sessions attended), number of adverse events during the study period, dropout and loss to follow-up, and description of the control group, if any. In addition, primary and secondary outcomes measured at baseline and following the exercise intervention will be extracted (mean and a measure of variance such as standard deviation [SD], standard error, 95% CI) including CRF, AF symptom frequency and severity, general and AF-specific QoL, resting blood pressure and heart rate, BMI, body composition, functional capacity, physical activity levels and anxiety and depression severity. Whenever possible, results from an intention to treat analysis will be used. Data will be extracted by a reviewer and verified by a second reviewer. Any disagreements between the reviewers will be resolved by consensus and/or discussion with a third reviewer (JLR). Study authors will be contacted to resolve any uncertainties and for missing information. If studies include both sexes but fail to report outcomes by sex, the corresponding authors will be contacted to obtain sex-specific data. The approach to contact authors will involve up to 3 emails or 3 phone calls (59). Receptive authors will be provided a pre-specified data extraction form and will receive up to 3 follow-up emails or phone calls as reminders, after which they will be considered unwilling or unable to share their data. Authors of studies which report on collecting a measure of interest but do not report the outcome on the manuscript will be contacted to solicit the unpublished data using the same approach.

### Methodological quality and risk of bias within studies

For the assessment of study quality and risk of bias, standardized data forms will be completed by a reviewer (SVA or IRM) and verified by a second (SVA or IRM). Any disagreements will be resolved by consensus and/or discussion with a third reviewer (JLR). Reviewers will not be blinded to the authors or journals.

The TESTEX scale will be used to examine the quality and risk of bias of each individual study as it is a valid and reliable tool to assess the quality and reporting of exercise training trials (60). The TESTEX scale uses 12 criteria (some of which can score more than one point) for a maximum score of 15 points, 5 pertain to *study quality*: (1) eligibility criteria specified, (2) randomization specified, (3) allocation concealment, (4) groups similar at baseline and (5) blinding of assessor for the primary outcome; and 10 pertain to *study reporting*: (1) outcome measures assessed in 85% of patients, (2) intention-to-treat analysis, (3) between-group statistical comparisons reported, (4) point measures and measures of variability for all reported outcome measures, (5) activity monitoring in control groups, (6) relative exercise intensity remained constant and (7) exercise volume and energy expenditure.

### Assessment of overall quality of the evidence

The quality of the evidence for the included studies will be evaluated using the Grading of Recommendations Assessment, Development and Evaluation (GRADE) (61) approach. GRADE classifies the evidence as high, moderate, low or very low depending on: (1) risk of bias within individual studies, (2) inconsistency of results between studies, (3) indirectness of evidence, (4) imprecision and (5) publication bias. RCT evidence represents the highest quality rating.

### Quantitative data synthesis and planned analyses

#### Measures of treatment effect

If studies are sufficiently homogeneous regarding population, interventions, outcomes and study design, and contain a number of samples that would provide meaningful summary measures, meta-analyses will be performed. Continuous outcomes will be analysed using MDs, or standardized mean differences (SMD) if the measurement methods (e.g. scales) for the same outcome vary between studies. Categorical data will be presented descriptively. The MD will be determined by calculating the difference in the mean outcome between sexes and dividing by the SD of the outcome among participants.

Pooled estimates of the effect of exercise training on CRF for females when compared to males (i.e. the MD or SMD between the CRF changes in females vs. males following exercise training) will be obtained using a random-effects model, since effect sizes are not expected to be identical across all studies. The same approach will be followed for the secondary outcomes. Where possible, pooled estimates will be calculated for the effect of exercise training vs. usual care on CRF in each sex separately. A quantitative synthesis of the measures of effect and 95% CI will be presented in the form of forest plots, which will be created using the Cochrane RevMan 5.4.1 or Comprehensive Meta-Analysis software. If quantitative synthesis is not appropriate, a narrative synthesis will be presented following the Synthesis Without Meta-analysis (SWiM) guidelines (62).

#### Pre-specified additional analyses

Several sub-analyses including sensitivity and meta-regression approaches will be performed, if sufficient data are available, to elicit whether specific driving forces could influence different effect sizes. Such sub-analyses may elucidate factors and mechanisms influencing sex-based differences in the response to exercise and whether a specific exercise prescription maximizes the health benefits in females or males with AF. For instance, we plan to examine whether the:

(1) Exercise program characteristics (FITT) influence the change in CRF in females or males with AF;

(2) Participant baseline characteristics (e.g. sociodemographic and gender-related factors, physical- and mental-health) influence the change in CRF in females or males with AF;

(3) Method of CRF measurement (estimated vs. measured) influences the difference in change in CRF between females and males with AF;

(4) Sex-based differences in adherence influence sex differences in the change in CRF.

#### Assessment of heterogeneity and publication bias

The degree of inconsistency across studies will be quantified using the I^2^ statistic, which describes the percentage of total variation across studies that is due to heterogeneity rather than chance. I^2^ values of 25%, 50% and 75% have been classified as low, moderate and high heterogeneity (63). Where meta-analyses are performed, funnel plots will be prepared to assess the effect of publication bias on our systematic review. Visual inspection will be used to determine whether bias is absent (plots resemble a symmetrical inverted funnel) or present (skewed and asymmetrical shape). Further, an Egger’s test p<0.10 will indicate the presence of a significant publication bias (64).

We will take into consideration the methodological quality, risk of bias, heterogeneity and overall quality of the evidence when interpreting the treatment effect reported in our systematic review as they can influence the true effect of the exercise intervention and the clinical relevance of our findings.

## Supporting information

Additional File 1. PRISMA-P checklist.

Additional File 2. Draft of the data extraction form.

## Data Availability

Upon completion of this systematic review, the datasets used and/or analysed will be made available from the corresponding author on reasonable request.

## CLINICAL RELEVANCE

The lack of outcome reporting by sex in exercise studies may lead to suboptimal exercise volumes and health benefits in females and males with AF. This systematic review will address the lack of sex-based analyses in previously conducted exercise studies; and, provide evidence applicable to both sexes of the physical and mental health effects of exercise training in the AF population. Whether females and males with AF respond differently to exercise is unknown, but the notable clinical sex-based differences in AF warrant sex consideration when developing exercise recommendations. Our findings will be of value to patients with AF, health researchers and healthcare providers involved in AF management.

### DISSEMINATION

The findings from this systematic review will be disseminated through peer-reviewed publications and conference presentations/proceedings. Further, the authors will invite health researchers, patients with AF and healthcare providers involved in AF management to educational webinars focused on our findings. These webinars are expected to have global reach and equip patients and exercise specialists with information on exercise participation and prescription.

#### LIST OF ABBREVIATIONS

AF: Atrial Fibrillation
AFEQT: Atrial Fibrillation Effect on Quality-of-Life Scale
AFQLQ: Atrial Fibrillation Quality of Life Questionnaire
AF-QoL: Atrial Fibrillation Quality of Life
AFSS: University of Toronto Atrial Fibrillation Severity Scale
AMSTASR: Measurement Tool to Assess Systematic Reviews
ASTA: Arrhythmia-Specific Questionnaire in Tachycardia and Arrhythmia
BAI: Beck anxiety inventory
BDI: Beck Depression Inventory
BMI: Body Mass Index
CCS-SAF: Canadian Cardiovascular Society Severity in Atrial Fibrillation Scale
CDS: Cardiac Depression Scale
CRF: Cardiorespiratory Fitness
EHRA: European Heart Rhythm Association
EQ-5D: EuroQoL
FITT: Exercise program characteristics (frequency, intensity, time, type)
GAD-7: Generalized Anxiety Disorder
GRADE: Grading of Recommendations Assessment, Development and Evaluation
HADS: Hospital Anxiety and Depression Scale
MAFSI: Mayo Atrial Fibrillation Symptom Inventory
MD: Mean Difference
METs: Metabolic Equivalents
PHQ-9: Patient Health Questionnaire-9
PRISMA: Preferred Reporting Items for Systematic Review and Meta-Analyses
PROSPERO: International Prospective Register of Systematic Reviews
QLAF: Quality of Life in Atrial Fibrillation
QoL: Quality of Life
RCT: Randomized Controlled Trial
SCL: Arrhythmia Symptom Checklist Frequency and Severity
SD: Standard Deviation
SF-12/36: Medical Outcome Study Short-form Health Survey
SMD: Standardized Mean Difference
STAI: State-trait anxiety inventory
VO2peak: Peak Oxygen Consumption

## DECLARATIONS

### Authors’ contributions

SVA and JLR were responsible for the design of the systematic review protocol. SVA drafted the manuscript. IRM drafted the abstract. All authors critically revised the manuscript and gave final approval.

---

**Additional File 1**. PRISMA-P checklist.

**Additional File 2**. Draft of the data extraction form.

## References

1. Dai H, Zhang Q, Much AA, Maor E, Segev A, Beinart R, et al. Global, regional, and national prevalence, incidence, mortality, and risk factors for atrial fibrillation, 1990–2017: results from the Global Burden of Disease Study 2017. Eur Heart J Qual Care Clin Outcomes. 2021;7(6):574–582. doi: 10.1093/ehjqcco/qcaa061.

2. Lippi G, Sanchis-Gomar F, Cervellin G. Global epidemiology of atrial fibrillation: An increasing epidemic and public health challenge. International Journal of Stroke. 2021;16(2):217–21.

3. Padfield GJ, Steinberg C, Swampillai J, Qian H, Connolly SJ, Dorian P, et al. Progression of paroxysmal to persistent atrial fibrillation: 10-year follow-up in the Canadian Registry of Atrial Fibrillation. Heart Rhythm. 2017;14(6):801–7.

4. Andrade JG, Aguilar M, Atzema C, Bell A, Cairns JA, Cheung CC, et al. The 2020 Canadian Cardiovascular Society/Canadian Heart Rhythm Society Comprehensive Guidelines for the Management of Atrial Fibrillation. Canadian Journal of Cardiology. 2020.36(12):1847–948.

5. Buckley BJR, Harrison SL, Fazio-Eynullayeva E, Underhill P, Lane DA, Thijssen DHJ, et al. Association of Exercise-Based Cardiac Rehabilitation with Progression of Paroxysmal to Sustained Atrial Fibrillation. JCM. 2021;10(3):435.

6. Borland M, Bergfeldt L, Nordeman L, Bollano E, Andersson L, Rosenkvist A, et al. Exercise-based cardiac rehabilitation improves physical fitness in patients with permanent atrial fibrillation – a randomised controlled study. Translational Sports Medicine. 2020;3:415–425. doi: 10.1002/tsm2.166.

7. Hegbom F, Sire S, Heldal M, Orning OM, Stavem K, Gjesdal K. Short-term exercise training in patients with chronic atrial fibrillation: effects on exercise capacity, AV conduction, and quality of life. J Cardiopulm Rehabil. 2006;26(1):24–9.

8. Joensen AM, Dinesen PT, Svendsen LT, Hoejbjerg TK, Fjerbaek A, Andreasen J, et al. Effect of patient education and physical training on quality of life and physical exercise capacity in patients with paroxysmal or persistent atrial fibrillation: A randomized study. J Rehabil Med. 2019;51(6):442–450. doi: 10.2340/16501977-2551.

9. Kato M, Ogano M, Mori Y, Kochi K, Morimoto D, Kito K, et al. Exercise-based cardiac rehabilitation for patients with catheter ablation for persistent atrial fibrillation: A randomized controlled clinical trial. Eur J Prev Cardiolog. 2019;2047487319859974.

10. Malmo V, Nes BM, Amundsen BH, Tjonna A-E, Stoylen A, Rossvoll O, et al. Aerobic Interval Training Reduces the Burden of Atrial Fibrillation in the Short Term: A Randomized Trial. Circulation. 2016;133(5):466–73.

11. Mertens DJ, Kavanagh T. Exercise Training for Patients With Chronic Atrial Fibrillation. Journal of Cardiopulmonary Rehabilitation and Prevention. 1996;16(3):193.

12. Osbak PS, Mourier M, Kjaer A, Henriksen JH, Kofoed KF, Jensen GB. A randomized study of the effects of exercise training on patients with atrial fibrillation. American Heart Journal. 2011;162(6):1080–7.

13. Plisiene J, Blumberg A, Haager G, Knackstedt C, Latsch J, Norra C, et al. Moderate physical exercise: a simplified approach for ventricular rate control in older patients with atrial fibrillation. Clin Res Cardiol. 2008;97(11):820.

14. Risom SS, Zwisler A-D, Rasmussen TB, Sibilitz KL, Madsen TLS, Svendsen JH, et al. Cardiac rehabilitation versus usual care for patients treated with catheter ablation for atrial fibrillation: Results of the randomized CopenHeartRFA trial. American Heart Journal. 2016;181:120–9.

15. Skielboe AK, Bandholm TQ, Hakmann S, Mourier M, Kallemose T, Dixen U. Cardiovascular exercise and burden of arrhythmia in patients with atrial fibrillation - A randomized controlled trial. PLoS One. 2017;12(2).

16. Vanhees L, Schepers D, Defoor J, Brusselle S, Tchursh N, Fagard R. Exercise performance and training in cardiac patients with atrial fibrillation. J Cardiopulm Rehabil. 2000;20(6):346–52.

17. Pathak RK, Elliott A, Middeldorp ME, Meredith M, Mehta AB, Mahajan R, et al. Impact of CARDIOrespiratory FITness on Arrhythmia Recurrence in Obese Individuals With Atrial Fibrillation: The CARDIO-FIT Study. Journal of the American College of Cardiology. 2015;66(9):985–96.

18. Keteyian SJ, Brawner CA, Savage PD, Ehrman JK, Schairer J, Divine G, et al. Peak aerobic capacity predicts prognosis in patients with coronary heart disease. American Heart Journal. 2008;156(2):292–300.

19. Tsuneoka H, Koike A, Nagayama O, Sakurada K, Kato J, Sato A, et al. Prognostic Value of Cardiopulmonary Exercise Testing in Cardiac Patients With Atrial Fibrillation. Int Heart J. 2012;53(2):102–7.

20. Ariansena I, Gjesdala K, Abdelnoorb M, Edvardsenc E, Engerd S, Tveitd A. Quality of Life, Exercise Capacity and Comorbidity in Old Patients with Permanent Atrial Fibrillation. J Atr Fibrillation. 2008;1(4).

21. Elliott AD, Verdicchio CV, Gallagher C, Linz D, Mahajan R, Mishima R, et al. Factors Contributing to Exercise Intolerance in Patients With Atrial Fibrillation. Heart, Lung and Circulation. 2021;30(7):947–54.

22. Younis A, Shaviv E, Nof E, Israel A, Berkovitch A, Goldenberg I, et al. The role and outcome of cardiac rehabilitation program in patients with atrial fibrillation. Clin Cardiol. 2018;41(9):1170–6.

23. Wagner MK, Zwisler A-DO, Risom SS, Svendsen JH, Christensen AV, Berg SK. Sex differences in health status and rehabilitation outcomes in patients with atrial fibrillation treated with ablation: Results from the CopenHeartRFA trial. Eur J Cardiovasc Nurs. 2018;17(2):123–35.

24. Ball J, Carrington MJ, Wood KA, Stewart S, Investigators the S. Women Versus Men with Chronic Atrial Fibrillation: Insights from the Standard Versus Atrial Fibrillation spEcific managemenT studY (SAFETY). PLOS ONE. 2013;8(5):e65795.

25. Schnabel RB, Pecen L, Ojeda FM, Lucerna M, Rzayeva N, Blankenberg S, et al. Gender differences in clinical presentation and 1-year outcomes in atrial fibrillation. Heart. 2017;103(13):1024–30.

26. Blum S, Muff C, Aeschbacher S, Ammann P, Erne P, Moschovitis G, et al. Prospective Assessment of Sex□Related Differences in Symptom Status and Health Perception Among Patients With Atrial Fibrillation. J Am Heart Assoc. 2017;6(7).

27. Lip GYH, Laroche C, Boriani G, Cimaglia P, Dan G-A, Santini M, et al. Sex-related differences in presentation, treatment, and outcome of patients with atrial fibrillation in Europe: a report from the Euro Observational Research Programme Pilot survey on Atrial Fibrillation. Europace. 2015;17(1):24–31.

28. Piccini JP, Simon DN, Steinberg BA, Thomas L, Allen LA, Fonarow GC, et al. Differences in Clinical and Functional Outcomes of Atrial Fibrillation in Women and Men: Two-Year Results From the ORBIT-AF Registry. JAMA Cardiol. 2016;1(3):282–91.

29. Strømnes LA, Ree H, Gjesdal K, Ariansen I. Sex Differences in Quality of Life in Patients With Atrial Fibrillation: A Systematic Review. J Am Heart Assoc. 2019;8(8):e010992.

30. Diaz-Canestro C, Montero D. Sex Dimorphism of VO2max Trainability: A Systematic Review and Meta-analysis. Sports Med. 2019;49, 1949–1956. https://doi.org/10.1007/s40279-019-01180-z

31. Parker BA, Kalasky MJ, Proctor DN. Evidence for sex differences in cardiovascular aging and adaptive responses to physical activity. Eur J Appl Physiol. 2010;110(2):235–46.

32. Witvrouwen I, Van Craenenbroeck EM, Abreu A, Moholdt T, Kränkel N. Exercise training in women with cardiovascular disease: Differential response and barriers – review and perspective. Eur J Prev Cardiolog. 2019;2047487319838221.

33. Sandercock G, Hurtado V, Cardoso F. Changes in cardiorespiratory fitness in cardiac rehabilitation patients: A meta-analysis. International Journal of Cardiology. 2013;167(3):894–902.

34. Rengo JL, Khadanga S, Savage PD, Ades PA. Response to Exercise Training During Cardiac Rehabilitation Differs by Sex. J Cardiopulm Rehabil Prev. 2020;40(5):319–324. doi: 10.1097/HCR.0000000000000536.

35. Tannenbaum C, Norris CM, McMurtry MS. Sex-Specific Considerations in Guidelines Generation and Application. Canadian Journal of Cardiology. 2019;35(5):598–605.

36. Hegbom F, Stavem K, Sire S, Heldal M, Orning OM, Gjesdal K. Effects of short-term exercise training on symptoms and quality of life in patients with chronic atrial fibrillation. International Journal of Cardiology. 2007;116(1):86–92.

37. Rienstra M, Van Veldhuisen DJ, Hagens VE, Ranchor AV, Veeger NJGM, Crijns HJGM, et al. Gender-Related Differences in Rhythm Control Treatment in Persistent Atrial Fibrillation: Data of the Rate Control Versus Electrical Cardioversion (RACE) Study. Journal of the American College of Cardiology. 2005;46(7):1298–306.

38. Humphries Karin H., Kerr Charles R., Connolly Stuart J., Klein George, Boone John A., Green Martin, et al. New-Onset Atrial Fibrillation. Circulation. 2001;103(19):2365–70.

39. Lau Dennis H., Nattel Stanley, Kalman Jonathan M, Sanders Prashanthan. Modifiable Risk Factors and Atrial Fibrillation. Circulation. 2017;136(6):583–96.

40. Penedo FJ, Dahn JR. Exercise and well-being: a review of mental and physical health benefits associated with physical activity. Current Opinion in Psychiatry. 2005;18(2):189.

41. Risom SS, Zwisler A-D, Johansen PP, Sibilitz KL, Lindschou J, Gluud C, et al. Exercise□based cardiac rehabilitation for adults with atrial fibrillation. Cochrane Database of Systematic Reviews. 2017;2(2):CD011197. doi: 10.1002/14651858.CD011197.pub2

42. Mills M, Johnson E, Zafar H, Horwood A, Lax N, Charlesworth S, et al. An exercise-based cardiac rehabilitation programme for AF patients in the NHS: a feasibility study. The British Journal of Cardiology. 2020;27:67–70. doi:10.5837/bjc.2020.020.

43. Reed JL, Clarke AE, Faraz AM, Birnie DH, Tulloch HE, Reid RD, et al. The Impact of Cardiac Rehabilitation on Mental and Physical Health in Patients With Atrial Fibrillation: A Matched Case-Control Study. Canadian Journal of Cardiology. 2018;34(11):1512–21.

44. Trachsel LD, Boidin M, Henri C, Fortier A, Lalongé J, Juneau M, et al. Women and men with coronary heart disease respond similarly to different aerobic exercise training modalities: a pooled analysis of prospective randomized trials. Appl Physiol Nutr Metab. 2021;46(5):417–425. doi: 10.1139/apnm-2020-0650.

45. Way KL, Vidal-Almela S, Moholdt T, Currie KD, Aksetøy I-LA, Boidin M, et al. Sex Differences in Cardiometabolic Health Indicators after HIIT in Patients with Coronary Artery Disease. Medicine & Science in Sports & Exercise. 2021;53(7):1345–55.

46. Hindricks G, Potpara T, Dagres N, Arbelo E, Bax JJ, Blomström-Lundqvist C, et al. 2020 ESC Guidelines for the diagnosis and management of atrial fibrillation developed in collaboration with the European Association of Cardio-Thoracic Surgery (EACTS)The Task Force for the diagnosis and management of atrial fibrillation of the European Society of Cardiology (ESC) Developed with the special contribution of the European Heart Rhythm Association (EHRA) of the ESC. Eur Heart J. 2021;42(5):373–498. doi: 10.1093/eurheartj/ehaa612.

47. Chung MK, Eckhardt LL, Chen LY, Ahmed HM, Gopinathannair R, Joglar JA, et al. Lifestyle and Risk Factor Modification for Reduction of Atrial Fibrillation: A Scientific Statement From the American Heart Association. Circulation. 2020;141(16).

48. Reed JL, Terada T, Way KL, Vidal-Almela S, Pipe AL, Risom SS. Exercise Targets in the 2020 CCS Guidelines for the Management of Patients with Atrial Fibrillation. Canadian Journal of Cardiology. 2021;37(10):1678–1679. doi: 10.1016/j.cjca.2021.04.019

49. Shamseer L, Moher D, Clarke M, Ghersi D, Liberati A, Petticrew M, et al. Preferred reporting items for systematic review and meta-analysis protocols (PRISMA-P) 2015: elaboration and explanation. BMJ. 2015;349:g7647.

50. Page MJ, McKenzie JE, Bossuyt PM, Boutron I, Hoffmann TC, Mulrow CD, et al. The PRISMA 2020 statement: an updated guideline for reporting systematic reviews. BMJ. 2021;372:71.

51. Shea BJ, Reeves BC, Wells G, Thuku M, Hamel C, Moran J, et al. AMSTAR 2: a critical appraisal tool for systematic reviews that include randomised or non-randomised studies of healthcare interventions, or both. BMJ. 2017;j4008.

52. Hannan AL, Hing W, Simas V, Climstein M, Coombes JS, Jayasinghe R, et al. High-intensity interval training versus moderate-intensity continuous training within cardiac rehabilitation: a systematic review and meta-analysis. Open Access J Sports Med. 2018;9:1–17.

53. Caspersen CJ, Powell KE, Christenson GM. Physical activity, exercise, and physical fitness: definitions and distinctions for health-related research. Public Health Rep. 1985;100(2):126–31.

54. American College of Sports Medicine. ACSM’s Guidelines for Exercise Testing and Prescription, 11th ed. Lippincott Williams & Wilkins; 2020.

55. Reed JL, Cotie LM, Cole CA, Harris J, Moran B, Scott K, et al. Submaximal Exercise Testing in Cardiovascular Rehabilitation Settings (BEST Study). Front Physiol. 2020;10:1517.

56. Aliot E, Botto GL, Crijns HJ, Kirchhof P. Quality of life in patients with atrial fibrillation: how to assess it and how to improve it. Europace. 2014;16(6):787–96.

57. Steinberg BA, Dorian P, Anstrom KJ, Hess R, Mark DB, Noseworthy PA, et al. Patient-Reported Outcomes in Atrial Fibrillation Research: Results of a Clinicaltrials.gov Analysis. JACC: Clinical Electrophysiology. 2019;5(5):599–605.

58. Reed JL, Terada T, Chirico D, Prince SA, Pipe AL. The Effects of Cardiac Rehabilitation in Patients with Atrial Fibrillation: A Systematic Review. Canadian Journal of Cardiology 2018;34(10 Suppl 2):S284–S295. doi: 10.1016/j.cjca.2018.07.014.

59. Young T, Hopewell S. Methods for obtaining unpublished data. Cochrane Database of Systematic Reviews. 2011(11):MR000027. doi: 10.1002/14651858.MR000027.pub2.

60. Smart NA, Waldron M, Ismail H, Giallauria F, Vigorito C, Cornelissen V, et al. Validation of a new tool for the assessment of study quality and reporting in exercise training studies: TESTEX. Int J Evid Based Healthc. 2015;13(1):9–18.

61. Guyatt GH, Oxman AD, Vist GE, Kunz R, Falck-Ytter Y, Alonso-Coello P, et al. GRADE: an emerging consensus on rating quality of evidence and strength of recommendations. BMJ. 2008;336(7650):924–6.

62. Campbell M, McKenzie JE, Sowden A, Katikireddi SV, Brennan SE, Ellis S, et al. Synthesis without meta-analysis (SWiM) in systematic reviews: reporting guideline. BMJ. 2020;6890.

63. Higgins JPT, Thompson SG, Deeks JJ, Altman DG. Measuring inconsistency in meta-analyses. BMJ. 2003;327(7414):557–60.

64. Egger M, Smith GD, Schneider M, Minder C. Bias in meta-analysis detected by a simple, graphical test. BMJ. 1997;315(7109):629–34.

