## Additional File 1. PRISMA-P checklist. for "Sex differences in the change in cardiorespiratory fitness and additional physical and mental health outcomes after exercise training in adults with atrial fibrillation: a systematic review protocol"

### PRISMA-P 2015 Checklist

This checklist has been adapted for use with systematic review protocol submissions to BioMed Central journals from Table 3 in Moher D et al: Preferred reporting items for systematic review and meta-analysis protocols (PRISMA-P) 2015 statement. *Systematic Reviews* 2015 **4**:1

An Editorial from the Editors-in-Chief of *Systematic Reviews* details why this checklist was adapted - Moher D, Stewart L & Shekelle P: Implementing PRISMA-P: recommendations for prospective authors. *Systematic Reviews* 2016 **5**:15

| Section/topic | # | Checklist item | Information reported | | Line number(s) |
|---|---|---|---|---|---|
| | | | Yes | No | |
| **ADMINISTRATIVE INFORMATION** | | | | | |
| **Title** | | | | | |
| Identification | 1a | Identify the report as a protocol of a systematic review | ☒ | ☐ | Title |
| Update | 1b | If the protocol is for an update of a previous systematic review, identify as such | ☐ | ☒ | N/A |
| **Registration** | 2 | If registered, provide the name of the registry (e.g., PROSPERO) and registration number in the Abstract | ☒ | ☐ | End of abstract |
| **Authors** | | | | | |
| Contact | 3a | Provide name, institutional affiliation, and e-mail address of all protocol authors; provide physical mailing address of corresponding author | ☒ | ☐ | Title page |
| Contributions | 3b | Describe contributions of protocol authors and identify the guarantor of the review | ☒ | ☐ | Authors' contributions section |
| **Amendments** | 4 | If the protocol represents an amendment of a previously completed or published protocol, identify as such and list changes; otherwise, state plan for documenting important protocol amendments | ☒ | ☐ | Page 8, L 6-8 |
| **Support** | | | | | |
| Sources | 5a | Indicate sources of financial or other support for the review | ☒ | ☐ | Funding section |
| Sponsor | 5b | Provide name for the review funder and/or sponsor | ☐ | ☒ | N/A |
| Role of sponsor/funder | 5c | Describe roles of funder(s), sponsor(s), and/or institution(s), if any, in developing the protocol | ☐ | ☒ | N/A |

BioMed Central
The Open Access Publisher

Sex differences in the change in cardiorespiratory fitness and additional physical and mental health outcomes after exercise training in adults with atrial fibrillation: a systematic review protocol

| Section/topic | # | Checklist item | Information reported Yes | No | Line number(s) |
|---|---|---|---|---|---|
| **INTRODUCTION** | | | | | |
| **Rationale** | 6 | Describe the rationale for the review in the context of what is already known | ☒ | ☐ | Pages 6 to 7 |
| **Objectives** | 7 | Provide an explicit statement of the question(s) the review will address with reference to participants, interventions, comparators, and outcomes (PICO) | ☒ | ☐ | Page 7, L 13-20 |
| **METHODS** | | | | | |
| **Eligibility criteria** | 8 | Specify the study characteristics (e.g., PICO, study design, setting, time frame) and report characteristics (e.g., years considered, language, publication status) to be used as criteria for eligibility for the review | ☒ | ☐ | Page 8, L10 to Page 10, L 21 |
| **Information sources** | 9 | Describe all intended information sources (e.g., electronic databases, contact with study authors, trial registers, or other grey literature sources) with planned dates of coverage | ☒ | ☐ | Page 10, L23 to page 11 L 11<br>Page 13, L1-8 |
| **Search strategy** | 10 | Present draft of search strategy to be used for at least one electronic database, including planned limits, such that it could be repeated | ☒ | ☐ | Table 1. |
| ***STUDY RECORDS*** | | | | | |
| Data management | 11a | Describe the mechanism(s) that will be used to manage records and data throughout the review | ☒ | ☐ | Page 11, L13-15<br>Page 12, L4-6 |
| Selection process | 11b | State the process that will be used for selecting studies (e.g., two independent reviewers) through each phase of the review (i.e., screening, eligibility, and inclusion in meta-analysis) | ☒ | ☐ | Page 11, L12 to Page 12, L2 |
| Data collection process | 11c | Describe planned method of extracting data from reports (e.g., piloting forms, done independently, in duplicate), any processes for obtaining and confirming data from investigators | ☒ | ☐ | Page 12, L3 to Page 13 L8 |
| **Data items** | 12 | List and define all variables for which data will be sought (e.g., PICO items, funding sources), any pre-planned data assumptions and simplifications | ☒ | ☐ | Page 12 L6 to 21 |
| **Outcomes and prioritization** | 13 | List and define all outcomes for which data will be sought, including prioritization of main and additional outcomes, with rationale | ☒ | ☐ | Page 12 L6 to 21 |
| **Risk of bias in individual studies** | 14 | Describe anticipated methods for assessing risk of bias of individual studies, including whether this will be done at the outcome or study level, or both; state how this information will be used in data | ☒ | ☐ | Page 13, L9 to page 14 L6 |

Sex differences in the change in cardiorespiratory fitness and additional physical and mental health outcomes after exercise training in adults with atrial fibrillation: a systematic review protocol

| Section/topic | # | Checklist item | Information reported Yes | Information reported No | Line number(s) |
|---|---|---|---|---|---|
| | | synthesis | | | |
| ***DATA*** | | | | | |
| **Synthesis** | 15a | Describe criteria under which study data will be quantitatively synthesized | ☒ | ☐ | Page 13, L8 to 15 |
| | 15b | If data are appropriate for quantitative synthesis, describe planned summary measures, methods of handling data, and methods of combining data from studies, including any planned exploration of consistency (e.g., $I^2$, Kendall's tau) | ☒ | ☐ | Page 13, L11 to 25<br><br>Page 15, L15 to Page 16, L2 |
| | 15c | Describe any proposed additional analyses (e.g., sensitivity or subgroup analyses, meta-regression) | ☒ | ☐ | Page 15, L1 to 14 |
| | 15d | If quantitative synthesis is not appropriate, describe the type of summary planned | ☒ | ☐ | Page 14, L 23 to 25 |
| **Meta-bias(es)** | 16 | Specify any planned assessment of meta-bias(es) (e.g., publication bias across studies, selective reporting within studies) | ☒ | ☐ | Page 15, L15 to Page 16, L2 |
| **Confidence in cumulative evidence** | 17 | Describe how the strength of the body of evidence will be assessed (e.g., GRADE) | ☒ | ☐ | Page 14, L1-6 |
